# Optimizing dosing strategies for children with leprosy: A weight-banded dosing approach

**DOI:** 10.64898/2026.11.11.26367824

**Authors:** Jennie van Dyk, Tiziana Masini, Saba M. Lambert, Joydeepa Darlong, Patricia D. Deps, Mauricio L. Nobre, Wim van Brakel, Becks Hill, Amar Bahadur Timalsina, Sushant Satish Mane, Parul Malik, Vivek Lal, Roeland E. Wasmann

**Affiliations:** Division of Clinical Pharmacology, Department of Medicine, University of Cape Town, Cape Town, South Africa; Global Accelerator for Paediatric Formulations, Science for Health Department, World Health Organization, Geneva, Switzerland; Faculty of Infectious and Tropical Diseases, London School of Hygiene and Tropical Medicine, London WC1E 7HT, UK; The Leprosy Mission Trust India, New Delhi, India; Faculty of Medicine, Department of Public Health, Postgraduate Program of Infectious Diseases, Federal University of Espirito Santo, Vitoria-ES, Brazil; Tropical Disease Institute, Federal University of Rio Grande do Norte State, Brazil; International Leprosy Association, Brentford, London TW8 0QH, UK; Director of Programmes and Advocacy at Lepra, Colchester, UK; IDEA International, Kathmandu, Nepal; Grant Government Medical College, Pediatric Pulmonology Division, Sir J.J. Group of Hospitals, Mumbai, India; Global Leprosy Programme, World Health Organization, Geneva, Switzerland

## Abstract

**Background:** Dosing guidance for children with leprosy remains operationally challenging. For children under 10 years of age, current WHO recommendations rely on mg/kg dosing of the three separate components of leprosy multidrug therapy, which are difficult to implement in routine care and may result in inconsistent drug exposures. Weight-banded dosing offers a pragmatic alternative that better aligns with pharmacokinetic principles and existing drug formulations.

**Methodology:** A pharmacokinetic simulation study was conducted to develop harmonized weight-banded dosing regimens for rifampicin, clofazimine, and dapsone in children aged 2 to 10 years. Using the WHO dosing tool, drug exposures across harmonized weight bands were simulated and adult-equivalent systemic exposure (80–125% of adult area under the concentration–time curve) targeted. Predicted exposures were contextualized against safety reference regimens used in tuberculosis meningitis, multidrug-resistant tuberculosis, and leprosy. An iterative review process involving the WHO Technical Working Group on leprosy in children ensured clinical and operational feasibility.

**Results:** Proposed weight-banded multidrug therapy regimens achieved adult-equivalent exposures across most weight bands. Predicted exposures modestly exceeded 125% of adult levels at the lower end of selected weight bands but remained well below safety benchmarks, namely exposures associated with daily rifampicin therapy for tuberculosis meningitis and clofazimine regimens for multidrug-resistant tuberculosis. Dapsone exposures were comparable to those achieved with current leprosy dosing. The existing 10–14-year-old child MDT blister pack could be used without modification in children weighing 25–39.9 kg and with limited tablet or gel capsule removal in children weighing 6–24.9 kg.

**Conclusions:** A harmonized weight-banded dosing strategy was developed that enables effective, practical dosing of leprosy multidrug therapy in children under 10 years using existing free-of-charge blister packs provided through WHO. This approach improves dosing consistency, supports programmatic implementation, and provides a rational interim solution while child-friendly formulations are being developed.

**Author Summary:** Leprosy continues to affect children in many low-resource settings, yet guidance on how to dose treatment in younger children remains limited. Current recommendations often require clinicians to calculate mg/kg doses, which can be difficult in practice and may not match the oral formulations that are available.

In this study, we used computer-based simulations to develop a simplified dosing approach for the three drugs used to treat leprosy. We showed that grouping children into weight bands, rather than calculating individual doses, can achieve drug exposures similar to those seen in adults, where treatment is known to be effective. Importantly, we found that these doses can be delivered using existing pre-packed treatment kits by removing selected dose units, making this approach practical in routine care.

This work provides a more straightforward and reliable way to treat children with leprosy, reducing the burden on clinicians and improving access to treatment. However, challenges remain, including the lack of child-friendly formulations and limited data on how these drugs behave in children. Further studies are needed to improve treatment for this vulnerable population.

## Introduction

Leprosy remains an important cause of morbidity in socially vulnerable communities across South-East Asia, South America, and Africa [1]. The condition is caused by *Mycobacterium leprae* and *Mycobacterium lepromatosis*, which primarily targets the peripheral nervous system and skin [2] and can cause irreversible neuropathy, visible deformities, and lifelong disability if diagnosis and treatment are delayed. Owing to its long incubation period, leprosy is typically observed in children after the age of eight, however, cases have been reported in children as young as two years old [3,4]. In 2024, around 10,000 new child cases were reported globally, with 2.7% of affected children presenting with permanent impairment [1].

The World Health Organization (WHO)-recommended standard multidrug therapy (MDT) with rifampicin, clofazimine, and dapsone has been highly effective in reducing the global prevalence of leprosy [1]. The adult regimen includes a monthly dose of 600 mg rifampicin and 300 mg clofazimine, and a daily dose of 100 mg dapsone and 50 mg clofazimine, with good clinical outcomes and low relapse rates [5].

WHO guidance for the management of children with leprosy is included in the *Global Leprosy Strategy 201c–2020 Operational Manual* [6] and the *2018 Guidelines for the Diagnosis, Treatment and Prevention of Leprosy* [7]. In children and young adolescents aged 10 to 14 years, the recommended MDT consists of monthly 450 mg rifampicin and 150 mg clofazimine, daily 50 mg dapsone, and 50 mg clofazimine every other day. WHO has facilitated the worldwide provision of leprosy MDT free-of-charge through a donation programme from The Nippon Foundation during 1995-1999 and from Novartis since 2000. Through this programme, two types of pre-packed blister packs – containing rifampicin capsules, clofazimine soft gel capsules, and dapsone tablets - are supplied to support treatment of adults and adolescents aged 10 to 14 years [6].

For children below 10 years of age or weighing below 40 kg, WHO recommends 10 mg/kg rifampicin and 100 mg clofazimine monthly, 2 mg/kg dapsone daily, and 50 mg clofazimine twice weekly [7]. The *2018 Guidelines for the Diagnosis, Treatment and Prevention of Leprosy* state that it would be possible to adapt the existing peadiatric blister pack for children weighing 20–40 kg by halving the daily dapsone tablet, taking clofazimine twice weekly, and using a single 300 mg rifampicin capsule [6,7]. However, these adaptations are difficult to implement, as all formulations must be swallowed whole and the dapsone tablet in the blister pack is not scored, limiting dose flexibility [8].

Since no specific MDT combination blister packs are available for this group, guidelines recommend treatment using standalone formulations of the individual drugs. This requires clinicians to calculate the final dose (in mg) and assemble regimens matching the final mg dose by using formulations they have available, which poses several challenges and may lead to dosing and acceptability issues. For instance, rifampicin is most commonly available as 150 mg and 300 mg capsules, which are not acceptable to younger children given swallowing difficulties. The formulation strength is also not appropriate to deliver the required dosing in this population. A 20 mg/mL oral liquid formulation exists, but it is not widely available or part of the donation program and has a different bioavailability than rifampicin capsules, potentially leading to dose errors [9]. Clofazimine is available as 50 mg and 100 mg solid oral dosage forms, including soft gel capsules and tablets. Clofazimine soft-gel capsules allow for limited dose flexibility and are not acceptable for children who have difficulty swallowing them whole. Unlike hard capsules, they cannot be opened, and the contents administered by dispersing them in water. Clofazimine tablets are not formulated as dispersible tablets, but they can be administered after being dispersed in water and have a better stability profile than capsules. Clofazimine tablets are available within National Tuberculosis Programmes through the Stop TB Partnership Global Drug Facility [10]. Dapsone is available only as solid oral tablets, commonly in strengths of 25 mg, 50 mg, and 100 mg. In some markets, these tablets have functional scoring lines allowing for lower dose increments, but acceptability for younger children remains low, as tablets are intended to be administered whole.

Collectively, these factors limit the feasibility of implementing mg/kg dosing in routine clinical settings. Pharmacokinetically, fixed mg/kg dosing can result in inconsistent exposures in children, systematically underexposing smaller individuals because it does not account for the nonlinear scaling of drug clearance with body size [11]. As a consequence, smaller children require relatively higher mg/kg doses than larger children to achieve target exposures. Weight-banded dosing is therefore increasingly recommended and used, including in WHO dosing guidance for several diseases, as it better aligns dosing with physiological differences and helps ensure adequate systemic exposures across pediatric weight ranges [12].

The 20th meeting of the WHO Technical Advisory Group (TAG) on Leprosy recognized the need to address these challenges and recommended the establishment of a working group to examine issues related to leprosy in children and provide recommendations to WHO on undertaking an optimization exercise for addressing existing gaps in treatment protocols. Following the recommendation of TAG-Leprosy, a Technical Working Group (TWG) was established by the WHO Global Leprosy Programme with the aim to review current practices for the diagnosis of leprosy in children, evaluate existing treatment protocols and recommend best practices for management of leprosy in children. We conducted a simulation study aimed at developing a harmonized weight-banded dosing strategy for rifampicin, clofazimine, and dapsone in children under 10 years of age. Because WHO-distributed free-of-charge MDT blister packs remain the most convenient approach for delivering leprosy treatment, the study had two primary objectives: to identify weight-banded pediatric doses of rifampicin, clofazimine, and dapsone for children under 10 years old that achieve adult-comparable exposures, and to determine if such dosing strategies could be implemented with the 10–14-year-old child MDT blister pack, despite acknowledging potential issues with acceptability and feasibility.

## Methodology

The WHO paediatric dosing tool [11] was used to simulate leprosy MDT doses across WHO harmonized paediatric weight bands [12]. The dosing tool predicts steady-state systemic exposure (area under concentration-time curve, AUC) relative to an adult reference, accounting for differences in body size and, where applicable, metabolic maturation. The goal was to identify paediatric doses expected to achieve adult-equivalent exposure, defined as 80–125% of the expected adult exposure, consistent with bioequivalence standards [13]. Our adult reference was a person weighing 60 kg receiving standard MDT (600 mg rifampicin once monthly, 300 mg clofazimine once monthly followed by 50 mg daily, and 100 mg dapsone daily) [7].

Because of the long incubation of leprosy and very rare occurrence of cases in children under 2 years old, our simulations focused on children above this age assuming that clearance mechanisms were fully matured for all three drugs in this population by this age [11,14,15].

To contextualize the proposed regimens in children, we simulated paediatric reference therapies as pragmatic safety benchmarks. For rifampicin, the once-monthly MDT dose was compared against daily weight-banded dosing used in tuberculosis meningitis (e.g. 300 mg/day for the 10 to <13 kg children) [16,17].

Clofazimine exposures were compared with weight-banded clofazimine doses used in second-line multi-drug-resistant tuberculosis (e.g. 50 mg three times weekly for children 5–<10 kg) [16]. Clofazimine has a long half-life of approximately 4 months in children [18], enabling extended dosing intervals across weight bands (e.g. daily, alternate-day, twice weekly doses). Total monthly exposure was used to compare regimens to account for different dosing schedules.

Dapsone was compared with 25 mg daily dapsone doses used in some pediatric leprosy protocols for children below 10 years of age [6]. Specific weight-banded doses of each safety reference standard are provided in Table S1. The resulting exposure ranges were used as safety reference standards against which exposures from the proposed regimens were evaluated.

Throughout the development of the optimized dosing guidance, the TWG provided clinical and operational perspectives on leprosy management in children, including field experience, potential adherence challenges, suitability of formulations for different age groups, and practical considerations such as the ability of children to swallow tablets. This feedback ensured that the pharmacokinetic simulations were compatible with real-world clinical practice and informed adjustments throughout the modeling process.

## Results

The relative exposures of the final weight-banded rifampicin, clofazimine, and dapsone MDT doses, along with safety reference regimens, were simulated using the WHO pediatric dosing tool and are visualized in Figure 1. Overall, the selected doses achieved 80-125% adult-equivalent exposures across most weight bands while remaining well within the upper exposure thresholds of each pediatric safety reference regimens.

**Figure 1.**
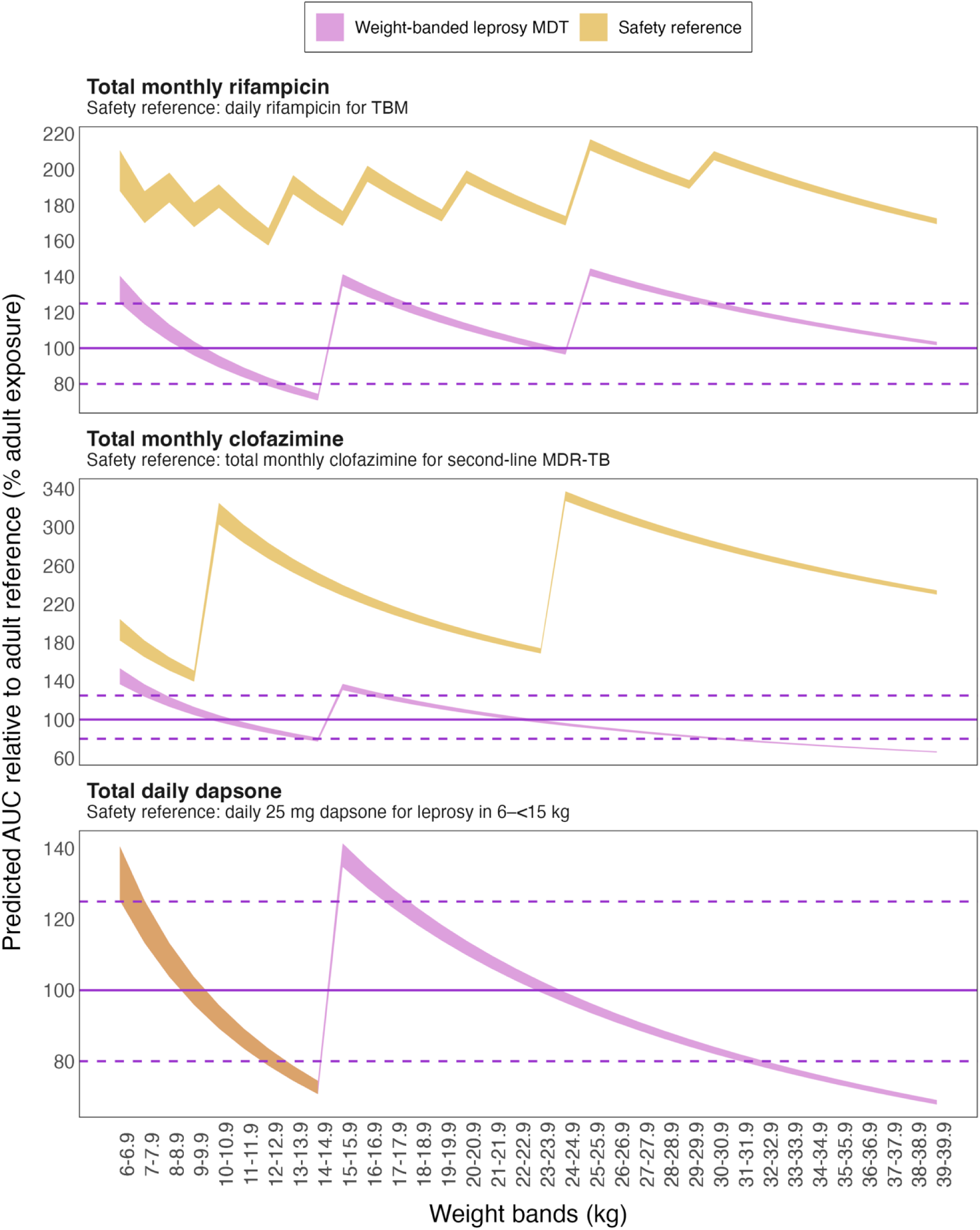
Simulated multidrug therapy (MDT) exposures by weight band in children aged >2 years and weighing 6–3G.G kg. Predicted adult-equivalent exposures (shaded area) for children receiving the proposed weight-banded MDT regimens for leprosy (purple) are shown alongside exposures predicted under safety reference standard therapies (orange). For dapsone, predicted exposures under the proposed regimen are equivalent to the safety reference exposure range, resulting in overlapping purple and orange distributions. The dotted line indicates 100% adult-equivalent exposure, and dashed lines indicate the 80–125% bioequivalence interval relative to adults receiving standard MDT for leprosy. TBM, tuberculosis meningitis; MDR-TB, multidrug resistant tuberculosis.

### Rifampicin exposure simulations

We evaluated weight-banded rifampicin doses for pediatric leprosy MDT, which are given once a month, and compared them with rifampicin administered once a day in children treated for tuberculosis meningitis as a conservative safety benchmark.

For the proposed weight-banded rifampicin doses, predicted exposures reached approximately 125–140% of adult-equivalent leprosy exposure at weights of 6–6.9 kg (150 mg once-monthly), 15–16.9 kg (300 mg once-monthly), and 25–29.9 kg (300 mg once-monthly). These values represent total exposure from a single monthly dose and are much lower than the daily rifampicin exposure given to children experiencing tuberculosis meningitis (approximately 160–220% of adult leprosy levels). Despite slightly exceeding the adult-equivalent range, the selected once-monthly rifampicin MDT doses were retained because they achieve equivalent exposure to standard adult MDT regimen while remaining well below the daily exposure margins observed in pediatric tuberculosis meningitis therapy.

Only in children weighing 13–14.9kg (150 mg once-monthly) is rifampicin exposure predicted to fall slightly below the adult-equivalence target, corresponding to approximately 75% of adult-equivalent exposure. This reduction was modest and did not justify escalation to a 300 mg monthly dose, which would result in exposures approaching tuberculosis meningitis daily exposure margins.

### Clofazimine exposure simulations

We evaluated weight-banded clofazimine doses for pediatric leprosy MDT against clofazimine doses administered to children as part of second-line therapy for multidrug-resistant tuberculosis, which served as the safety benchmark. Total monthly clofazimine exposures predicted under these multidrug-resistant tuberculosis dosing schedules reached approximately 140–340% of adult leprosy exposure, representing the upper exposure threshold in children.

Under the proposed weight-banded MDT regimen, clofazimine exposures were predicted to reach approximately 125–140% of adult-equivalent levels in children weighing 6–6.9 kg (100 mg monthly; 50 mg twice weekly) and 15–16.9 kg (150 mg monthly; 50 mg on alternate days). Although these values exceeded the 125% adult-equivalent target range, no weight band approached the pediatric safety reference threshold, and the doses were therefore retained.

In children weighing 30 kg and above (150mg monthly; 50mg on alternate days), clofazimine exposures were predicted to fall below 80% of adult-equivalent MDT levels and declined progressively with increasing weight. Doses were not increased, as these represent the highest pediatric doses prior to transition to the standard adult regimen and are equivalent to standard MDT in children above 10 years. The predicted exposures were therefore considered acceptable, as they match those achieved in children of the same weight currently receiving this treatment.

### Dapsone exposure simulations

Reference exposures for dapsone were defined using 25 mg once-daily dosing in children weighing 6–<15 kg, with predicted exposures ranging from approximately 75–140% of adult-equivalent leprosy dosing. Under the proposed regimen, predicted exposures were broadly comparable to this reference. Exposures reached approximately 125–140% of adult-equivalent levels at the lower end of the 6–9.9 kg (50 mg every other day) and 15–19.9 kg (50 mg daily) weight bands.

In children weighing 30 kg and above (50 mg daily), dapsone exposures were predicted to decline below 80% of adult-equivalent MDT levels and decreased progressively with increasing body weight. As with clofazimine, these doses are equivalent to standard MDT in children above 10 years. The lower predicted exposures are therefore consistent with those expected in children of the same weight currently receiving this treatment.

### Weight-banded leprosy MDT doses

Proposed weight-banded MDT doses of rifampicin, clofazimine, and dapsone, selected based on simulation results, are presented in Table 1, alongside a suggested guide for achieving these doses using the 10–14-year-old child MDT blister pack.

**Table 1.**
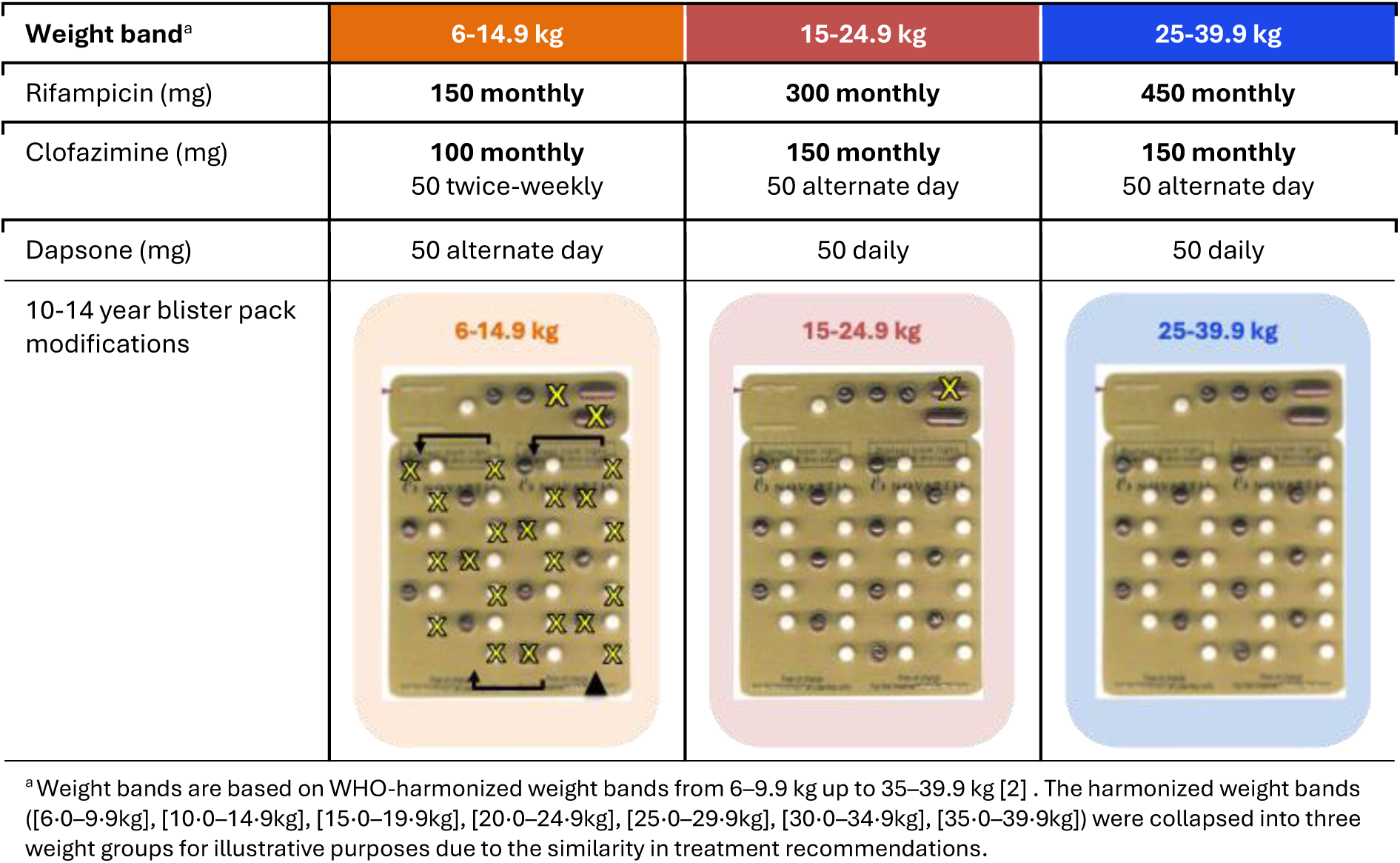
Weight-banded leprosy multidrug therapy (MDT) for children aged 2 to 10 years. Dosing table below shows monthly and daily doses of rifampicin, clofazimine, and dapsone across WHO-harmonized weight bands (6-39.9 kg). Suggested dose delivery using modified 10-14 year child blister packs is included. Crosses (×) indicate tablets to be removed by the healthcare worker prior to dispensing; children weighing 25–39.9 kg can use the blister pack without modifications.

| **Weight band<sup>a</sup>** | **6-14.9 kg** | **15-24.9 kg** | **25-39.9 kg** |
|---|---|---|---|
| Rifampicin (mg) | **150 monthly** | **300 monthly** | **450 monthly** |
| Clofazimine (mg) | **100 monthly**<br>50 twice-weekly | **150 monthly**<br>50 alternate day | **150 monthly**<br>50 alternate day |
| Dapsone (mg) | 50 alternate day | 50 daily | 50 daily |
| 10-14 year blister pack modifications | **6-14.9 kg** | **15-24.9 kg** | **25-39.9 kg** |

<sup>a</sup>Weight bands are based on WHO-harmonized weight bands from 6–9.9 kg up to 35–39.9 kg [2] . The harmonized weight bands ([6·0–9·9kg], [10·0–14·9kg], [15·0–19·9kg], [20·0–24·9kg], [25·0–29·9kg], [30·0–34·9kg], [35·0–39·9kg]) were collapsed into three weight groups for illustrative purposes due to the similarity in treatment recommendations.

Harmonized weight-banded dosing for children below 40 kg can be simplified into three practical regimens (6-14.9 kg, 15-24.9 kg, and 25-39.9 kg) for leprosy treatment with MDT. Children in the highest weight bands (25-39.9 kg) receive the standard MDT currently administered to 10–14-year-old children.

The existing 10–14-year-old child MDT blister pack could be used to deliver these doses as is in children weighing 25-39.9 kg and can be adjusted for those weighing under 25 kg by removing specific dose units. Only two scenarios of dose unit removal are required: for 15-24.9 kg children, the clinician may remove the 150 mg rifampicin monthly gel capsule from the blister pack. For individuals 6-14.9 kg, the 300 mg rifampicin monthly gel capsule must be removed, along with 1x 50 mg monthly clofazimine gel capsule, and then weekly dapsone tablets and clofazimine soft gel capsules as specified in the guide in Table 1.

## Discussion

Clinicians need clear and operationally feasible guidance for MDT dosing in young children with leprosy, where pharmacokinetic data are scarce or absent. In this study, we developed weight-banded dosing regimens for rifampicin, clofazimine, and dapsone using a model-informed approach. The proposed regimens are expected to achieve adult-equivalent exposures for standard MDT, providing a rational framework for dosing children with leprosy. Importantly, we found that these doses can be delivered using the existing child MDT blister pack in 2 to-10-year-old children after removing some dose units according to the weight-banded dosing.

The weight-banded dosing strategy can be implemented with minimal disruption to existing practice. Children weighing 25–39.9 kg can use the 10–14-year-old child blister pack as is, while those weighing 6–24.9 kg require clinician-guided removal of unnecessary tablets or capsules from the blister pack prior to distribution. This approach is both practical and accessible, given that MDT blister packs are distributed free-of-charge through national programs and other child-friendly single drug formulations are not yet readily available.

Modelling and simulation is recognized by major regulatory agencies as an essential tool in pediatric drug development to support evidence-based dosing recommendations [19,20]. Only a few studies have evaluated systemic exposures in children treated for leprosy, with little pharmacokinetic data available to assess dosing safety and efficacy [12]. Pediatric extrapolation from a reference (adult or other pediatric) population is an accepted regulatory approach to support evidence for safe and effective use of medicines in children, provided that the disease course and expected exposure–response are sufficiently similar between the target pediatric and reference populations [19,20]. Under these conditions, targeting adult-equivalent systemic exposures offers a reasonable surrogate target for pediatric dose selection.

Few pharmacokinetic studies have been conducted in leprosy, particularly in children [21]. In the absence of data, our simulations followed an accepted approach of extrapolation from reference populations. A 60-kg adult receiving standard MDT was chosen as the reference exposure to represent a typical adult body weight for endemic regions [22,23] and the acceptability range was defined based on EMA bioequivalence standards [13]. The acceptability range (in this case, adult equivalency) inevitably depends on the chosen adult reference; thus, predicted clofazimine and dapsone exposures below 80% of a 60 kg adult in children ≥30 kg do not necessarily indicate reduced efficacy. These doses are currently used in older (10–14-year-old) children receiving the blister pack as is and are assumed effective despite falling below adult-equivalent levels.

Our safety references for extrapolation were based on doses currently used in children. Rifampicin is well tolerated in children even at high daily doses used for tuberculosis meningitis [24], and the monthly exposures predicted here are well below those daily levels.

Prolonged use of clofazimine is associated with reversible hyperpigmentation but is otherwise generally well tolerated in children; even higher daily doses than those used in second-line pediatric tuberculosis regimens have minimal QTc prolongation [25], and our predicted exposures remained well below those benchmarks. Clofazimine-induced hyperpigmentation has been associated with increased social stigma, impaired social interaction, and reduced self-esteem [26]. Its impact on treatment adherence in children has not been quantified but should be considered during treatment monitoring.

Dapsone related adverse drug reactions are the most commonly reported in leprosy MDT [27], and there is limited dosing guidance outside of what is currently used in leprosy. Our predicted exposures are comparable to those expected in smaller (6-14.9 kg) children receiving a 25 mg daily dapsone regimen, which is generally assumed to be tolerated based on its use in existing leprosy protocols. Children in this weight band are typically younger than 3 years old, an age at which leprosy is statistically rare [3,4]. While malnutrition in some regions may contribute to older children falling within this weight band [28], tolerability data at this weight are based on very few cases.

Given the recognized risk of dapsone hypersensitivity reactions and haematological toxicity [29], clinical monitoring remains essential throughout the course of treatment, especially in the smallest children. Where feasible, baseline laboratory evaluation should be considered, including haemoglobin measurement and, ideally, full blood counts prior to treatment initiation, to support early detection of dapsone-associated anaemia and hypersensitivity reactions.

This work has some limitations. Predicted exposures represent population medians, and variability is expected across individual patients. While the average child is expected to achieve predicted exposure, clinical monitoring remains essential. The recommendations are intended for children over 2 years of age, as maturation effects in younger children require further investigation and better clinical data than what is currently available [21]. Given the long incubation period of leprosy [2], treatment under 2 years is unlikely [3,4], so these recommendations apply to the majority of children with leprosy. Our simulations assumed similar bioavailability between adults and children and does not provide clarity on the impact of comorbidities, such as malnutrition, on drug exposure [30]. Finally, although we illustrate how currently available MDT blister packs can be adapted to deliver the proposed doses in this study, we acknowledge that these formulations have important limitations for use in younger children as they were not designed to be acceptable to them. At present, however, they represent the best available option for delivering effective treatment free of charge. Future work should prioritize improving treatment acceptability by enabling access to child-friendly formulations that may already be available through other programmes (e.g. National Tuberculosis Programmes) and by fostering the development of child-friendly formulations, such as dispersible tablets, where they are not yet available, as has been highlighted in pediatric tuberculosis treatment [31].

This Pediatric Drug Optimization (PADO) process for childhood leprosy provides much greater confidence to clinicians managing these patients, rather than requiring them to calculate individual dosages that often do not align with available formulations. Adopting a weight-banded dosing approach allows existing MDT blister packs indicated for 10–14-year-olds to be used in children under 10 years to achieve adult-equivalent exposures. This provides an effective, free-of-charge treatment option for the youngest leprosy patients and delivers clear, practical dosing guidance for childhood leprosy.

In summary, the proposed weight-band dosing system represents a significant improvement in the management of leprosy treatment in young children. Acceptability of the drug formulations included in the MDT blister packs remains low for younger children and more work needs to be done to ensure that child-friendly formulations are available for children with leprosy. A PADO meeting for leprosy is planned by WHO to specifically investigate these issues [32]. Our work also highlights the scarcity of high-quality pharmacokinetic data in children with leprosy. Future clinical studies investigating pediatric leprosy should include pharmacokinetic evaluations to better understand drug exposures in this understudied population.

## Data Availability

All data produced in the present work are contained in the manuscript.

## Acknowledgements

We thank the members of the WHO Technical Working Group on Leprosy among Children for their expert guidance, clinical insights, and support throughout this work.

