## Supplementary material for "Optimizing dosing strategies for children with leprosy: A weight-banded dosing approach": table s1

### Supporting information

#### Table S1. Pediatric safety reference standard therapies.

| S1. Paediatric safety reference standard therapies | | | |
|---|---|---|---|
| Rifampicin [16, 17] | Weight band (kg) | Schedule description | Total dose (mg/day) |
| | 6–<8 | 75 mg × 3 | 225 mg |
| | 8–<10 | 75 mg × 3.5 | 262.5 mg |
| | 10–<13 | 75 mg × 4 | 300 mg |
| | 13–<16 | 75 mg × 5 | 375 mg |
| | 16–<20 | 75 mg × 6 | 450 mg |
| | 20–<25 | 75 mg × 7 | 525 mg |
| | 25–<30 | 75 mg × 9 | 675 mg |
| | 30–<35 | 75 mg × 10 | 750 mg |
| | 35–<50 | 150 mg × 4 | 600 mg |
| Clofazimine [16] | Weight band (kg) | Schedule description | Total dose (mg/month) |
| | 5–<7 | 50 mg M/W/F | 600 mg |
| | 7–<10 | 50 mg M/W/F | 600 mg |
| | 10–<16 | 50 mg daily | 1,400 mg |
| | 16–<24 | 50 mg daily | 1,400 mg |
| | 24–<30 | 100 mg daily | 2,800 mg |
| | 30–<36 | 100 mg daily | 2,800 mg |
| | 36–<46 | 100 mg daily | 2,800 mg |
| Dapsone [6] | Weight band (kg) | Total dose (mg/day) | |
| | 6–<15 | 25 mg | |

[6] World Health Organisation. Global leprosy strategy 2016–2020: accelerating towards a leprosy-free world – 2016 operational manual. New Delhi: Regional Office for South-East Asia; 2016.
[16] World Health Organization. Module 5: Management of tuberculosis in children and adolescents WHO operational handbook on tuberculosis. WHO operational handbook on tuberculosis, Geneva: 2022.
[17] Wasmann RE, Masini T, Viney K, Verkuijl S, Brands A, Hesseling AC, et al. A model-based approach for a practical dosing strategy for the short, intensive treatment regimen for tuberculous meningitis. Front Pharmacol 2023;14. https://doi.org/10.3389/fphar.2023.1055329.
